# PREDICTIVE IMMUNOLOGICAL, VIROLOGICAL, AND ROUTINE LABORATORY MARKERS FOR CRITICAL COVID-19 ON ADMISSION

**DOI:** 10.1101/2021.03.17.21253816

**Authors:** Immunocovid study, Mercedes García-Gasalla, Juana M Ferrer, Pablo A Fraile-Ribot, Adrián Ferre-Beltrán, Adrián Rodríguez, Natalia Martínez-Pomar, Luisa Ramon-Clar, Amanda Iglesias, Inés Losada-López, Francisco Fanjul, Joan Albert Pou, Isabel Llompart-Alabern, Nuria Toledo, Jaime Pons, Antonio Oliver, Melchor Riera, Javier Murillas

## Abstract

**Introduction:** Early identification of COVID-19 patients at risk of critical illness is challenging for clinicians. Immunological, virological, and routine laboratory markers to be used in addition to clinical data are needed.

**Aim and methods:** Blood tests to measure neutrophil/lymphocyte ratio (NLR), levels of ferritin, CRP, D-dimer, complement components (C3, C4), lymphocyte subsets, and cytokines, and SARS-Cov2 RT-PCR tests were performed in COVID-19 confirmed cases within 48 hours of admission. Cycle threshold (Ct) values were determined by RT-PCR from oral or nasopharyngeal swabs on the day of admission. Severity of symptoms was categorized as mild (grade 1), severe (grade 2), and critical (grade 3).

**Results:** 120 patients were included. COVID-19 was mild in 49, severe in 32, and critical in 39. Ferritin >370 ng/mL (OR 16.4, 95% CI 5.3-50.8), D-dimer >440 ng/mL (OR 5.45, 95% CI 2.36-12.61), CRP >7.65 mg/dL (OR 11.54, 95% CI 4.3-30.8), NLR >3.77 (OR 13.4, 95% CI 4.3-41.1), IL-6 >142.5 pg/mL (OR 8.76, 95% CI 3.56-21.54), IL-10 >10.8 pg/mL (OR 16.45, 95% CI 5.32-50.81), sIL-2rα (sCD25) >804.5 pg/mL (OR 14.06, 95% CI 4.56-43.28), IL-1Ra >88.4 pg/mL (OR 4.54, 95% CI 2.03-10.17), and IL-18 >144 pg/mL (OR 17.85, 95% CI 6.54-48.78) were associated with critical COVID-19 in the univariate age-adjusted analysis. In the multivariate age-adjusted analysis, this association was confirmed only for ferritin, CRP,NLR, IL-10, sIL-2rα, and IL-18. T, B, and NK cells were significantly decreased in critical patients. SARS-CoV-2 was undetected in blood except in 3 patients with indeterminate results. Ct values determined by RT-PCR from oral/nasopharyngeal swabs on admission were not related to symptom severity.

**Conclusion:** levels of ferritin, D-dimer, CRP, NLR, and cytokines and cytokine receptors IL-6, IL1-Ra, sCD25, IL-18, and IL-10, taken together with clinical data, can contribute to the early identification of critical COVID-19 patients.

## Background

The first cases of coronavirus disease 2019 (COVID-19) caused by the novel coronavirus 2 (SARS-CoV-2) were reported in China in December 2019 and rapidly escalated to a global pandemic with millions of confirmed cases worldwide, with numbers still rising [1-3]. Most COVID-19 cases are asymptomatic or result in only mild disease. However, in a substantial percentage of patients, respiratory illness requiring hospital care develops. Pulmonary disease can progress to critical illness with extensive lung damage and hypoxemic respiratory failure requiring prolonged ventilatory support [1]. Strategies for predicting the risk of disease progression are needed. Different clinical and laboratory markers to be used in the emergency setting are under investigation [4, 5], and several score models have been developed [6]. Identifying patients at high risk of critical illness on admission is challenging for clinicians, but is crucial for screening patients in need of more intensive treatment to improve prognosis. Several COVID-19 studies have shown an association between increased levels of serum inflammatory markers and proinflammatory cytokines and severe disease [7, 8]. A number of cytokines and chemokines have been implicated in the induction of the so-called cytokine storm [9-11] and some, notably interleukin (IL) 6, are now considered as both prognostic factors and therapeutic targets [12, 13]. Some investigators have also suggested a prognostic value of semi-quantitative real-time reverse transcription polymerase chain reaction (RT-PCR) from nasopharyngeal swabs, which determines viral load through cycle threshold (Ct) values, with lower Ct values associated with worse outcomes [14]. Moreover, SARS-CoV-2 viremia has been reported in severe cases [15].

The purpose of our study was to identify routine laboratory, immunological and virological biomarkers to be used together with clinical data for early identification of patients at risk of developing critical illness.

## Methods

### Participants and study design

Patients admitted to Son Espases and Son Llàtzer hospitals in Palma de Mallorca between 17 April and 20 July 2020 with positive nasopharyngeal swab RT-PCR test who accepted to participate in the study were included. Blood samples to measure the neutrophil/lymphocyte ratio (NLR), ferritin, C-reactive protein (CRP), D-dimer, complement components (C3, C4), lymphocyte subsets, and serum levels of cytokines, and to perform SARS-CoV-2-RT-PCR testing were obtained within the first 48 hours of admission.

The severity of signs and symptoms developed during hospitalization was categorized as mild (grade 1), severe (grade 2), and critical (grade 3). Mild disease was established when the patient had symptoms without pneumonia or had mild pneumonia; severe disease was established when dyspnea was associated with a respiratory rate ≥30/min or blood oxygen saturation <93%, or a partial pressure of arterial oxygen to fraction of inspired oxygen ratio <300, and/or lung infiltrates >50% within 24 to 48 hours from admission; critical disease was established for cases with respiratory failure, septic shock, and/or multiple organ dysfunction or failure [1]. The most severe category developed during hospitalization was selected, and it occurred in the first 72h of admission in all cases.

### Procedures

Routine blood examinations included leukocyte, neutrophil, and lymphocyte counts (cells*10^3/µL) and percentages. Serum biochemical tests recorded were ferritin (ng/L) determined by chemiluminescence immunoassay in architect i2000 equipment, CRP (mg/dL), and D-dimer (µg/L) quantified by immunoturbidimetry in architect 16.000, Immunological tests recorded were serum complement levels (C3, C4), lymphocyte subset cell counts (cells*10^3/µL) and percentages using flow cytometry, and plasma cytokine levels. We used an enzyme-linked immunosorbent assay (ELISA, DIAsource ImmunoAssays,SA, Belgium) to measure serum levels of IL-6, a chemiluminiscence assay (IMMULITE, Siemens, Germany) to determinate serum soluble IL-2 receptor alpha (sIL-2rα or sCD25), and a human cytokine magnetic bead panel (Merck Millipore, Billerica, MA, USA) to measure levels of other cytokines associated with cytokine storm: IL-1β, IL-1 receptor antagonist (IL-1Ra), IL-6, IL-8, IL-17A, IL-18, IL-22, interferon (IFN), tumor necrosis factor alpha (TNF-α), and IL-10.

SARS-CoV-2 was determined in nasopharyngeal swab specimens (within 16 hours of their collection) and in plasma samples stored at −70°C until testing. Nucleic acids were extracted using a Hamilton automated extraction platform and the amplification process was performed in a Bio-Rad CFX96 real-time PCR detection instrument (Bio-Rad, Hercules, CA) using two commercial RT-PCR kits: Allplex 2019-nCoV (Seegene, Seoul, Korea), which detects the presence of 3 target genes E gene, RdRP gene, and N gene; and LightMix® Modular SARS-CoV (COVID-19) E-gene (TIB MOLBIOL, Berlin, Germany).

In order to assess the correlation between semi-quantitative viral load values and pattern of disease severity, Ct values were recorded for each gene.

### Statistical analysis

Categorical variables were expressed as numbers and percentages, and continuous variables as mean and standard deviation (SD), or median and interquartile range (IQR) values. Proportions for categorical variables were compared using the χ2 test. The independent group t test and the Mann-Whitney U test were used for the comparison of continuous normally and not normally distributed variables, respectively. Normal distribution was studied by plotting histograms and with the Shapiro-Wilk test. A Kruskall Wallis test was performed to compare the difference between the three groups of patients classified according to disease severity. The receiver operating characteristic (ROC) curve was used to assess the diagnostic value of the biological markers, and the optimal cut-off value providing the best trade-off between sensitivity and specificity was selected with the Youden index. Univariate and multivariate age-adjusted logistic regression analyses were performed to explore the association between laboratory parameters and the risk of developing critical disease, using the values provided by the Youden index as cut-off points.

All statistical analyses were performed using SPSS (Statistical Package for the Social Sciences) version 22.0 software (SPSS Inc.). Two-sided *P* values of less than .05 were considered statistically significant.

## Results

Between 17 April and 20 July 2020, 120 laboratory-confirmed SARS-CoV-2 patients accepted to participate and were included in the study. There were 55 women and 65 men, and median age was 59 years (29-89). COVID-19 was considered mild in 49 patients, severe in 32 and critical in 39 cases. Twenty patients (16.67%) died during hospitalization. The death of 2 of them was not related to COVID-19 but to an associated malignancy. Pulmonary embolism (PE) was diagnosed in 15 patients (12.5%), and of these, 9 had critical COVID-19. Demographic characteristics and comorbidities of SARs-COV-2 infected patients with mild, severe, and critical disease are presented in Table 1. Compared with patients with mild or severe disease, critically ill patients were significantly more likely to be men (*p* = .00), have older age (*p* = .036), concomitant hypertension (*p* = .036), and diabetes mellitus (*p* = .023).

**Table 1.**
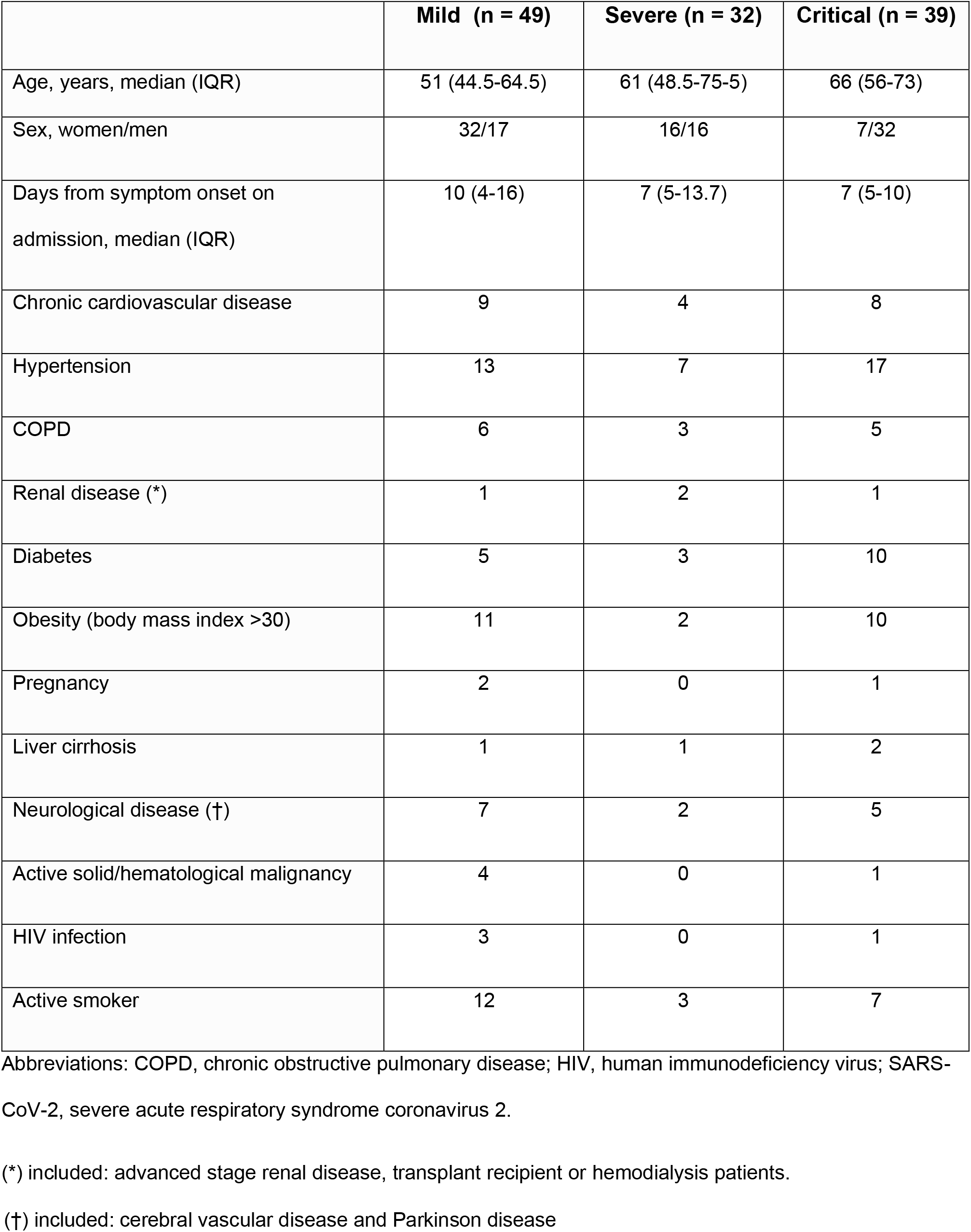
Demographic characteristics and comorbidities of SARs-COV-2 infected patients with mild, severe, and critical disease.

### Laboratory data

Laboratory markers were tested on admission (Table 2), and their cut-off values were calculated to predict the risk of developing critical COVID-19 (Fig 1). In the univariate analysis, ferritin > 370 ng/mL (OR [odds ratio] 16.4, 95% confidence interval [CI] 5.3-50.8), D-dimer >440 ng/mL (OR 5.45, 95% CI 2.36-12.61), CRP > 7.65 mg/dL (OR 11.54, 95% CI 4.3-30.8), and NLR > 3.77 (OR 13.4, 95% CI 4.3-41.1) predicted the development of critical COVID-19. In the multivariate analysis, the risk was statistically significant for ferritin (OR 8.1, 95% CI 2.1-30.6), NLR (OR 6.2, 95% CI 1.6-24.0), and CRP (OR 4.9, 95% CI 1.4-17.4), but not for D-dimer.

**Table 2.**
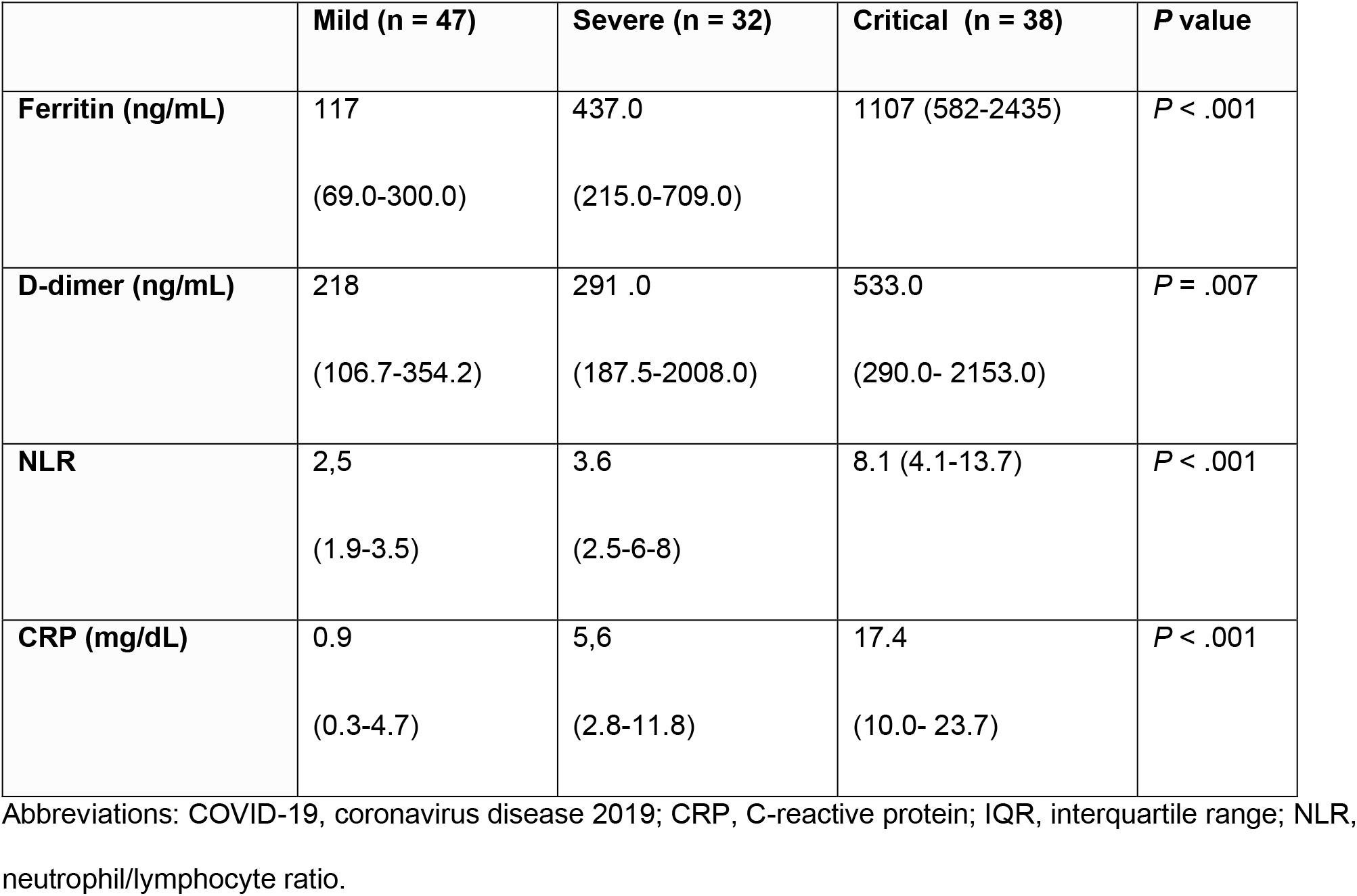
Comparison of laboratory markers in mild, severe, and critical COVID-19. Median and IQR.

**Figure 1.**
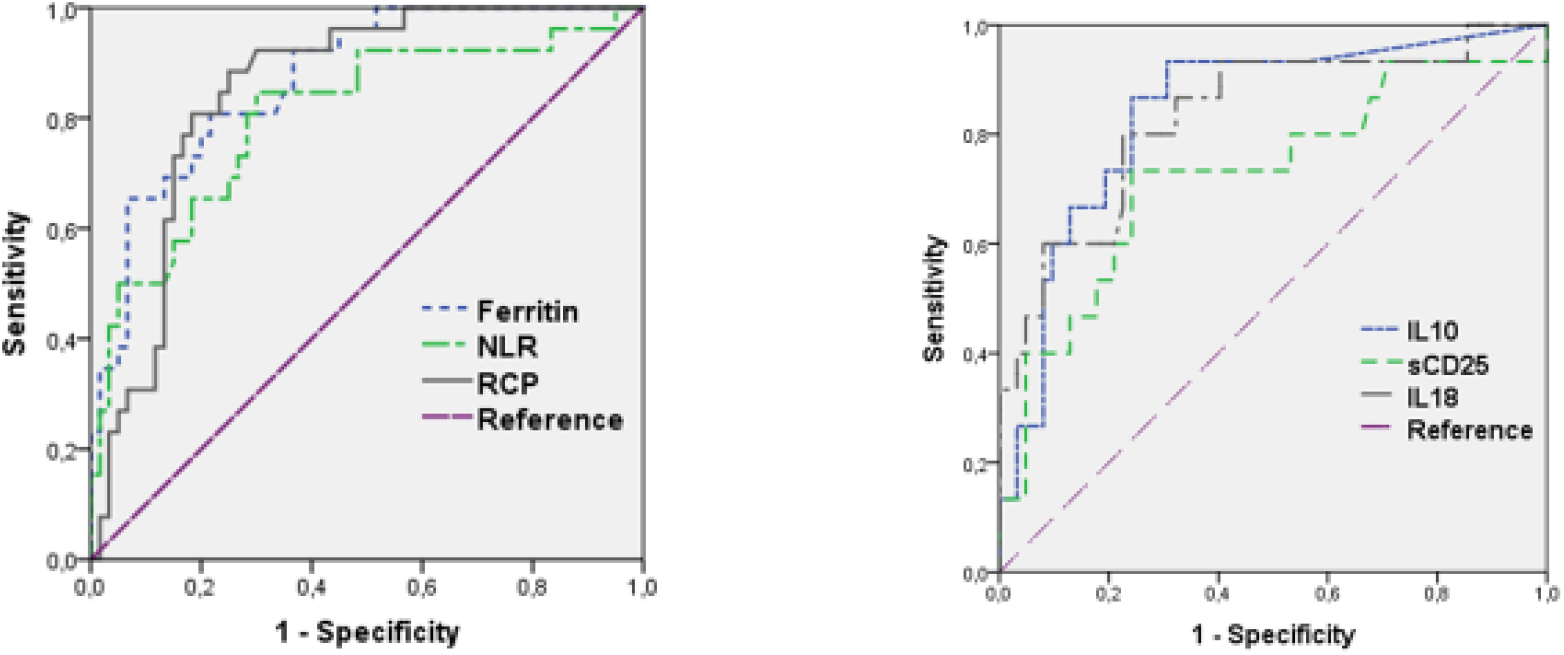
Performance of ROC curves in predicting critical patients for ferritin, CRP, NLR, IL-10, sCD25 (siL2rα), and IL-18. The univariateand multivariate logistic regression analysis distinguished critical from mild and severe disease patients Abbreviations: CRP, C-reactiv`e protein; NLR, neutrophil/lymphocyte ratio; s CD25s (sIL-2Rα) serum IL-2 receptor alpha.

### Immunological results

Serum complement levels were measured on admission. Median values of C3 and C4 levels in critical COVID-19 patients (group 3) were 116 mg/dL (108.0–127.7) and 26.0 mg/dL (17.0-39.0), respectively, whereas in mild and severe patients (groups 1 and 2) the median values were 125 mg/dL (105.5–145.5) and 26.5 mg/dL (21.0-33.0), respectively (*P* > .05). Low C4 levels (<20 mg/dL) were found in 5/15 (33.3%) patients with PE versus 22/105 (20.9%) patients without PE (*p* > .05) and in 10/39 (25.6%) patients with critical disease versus 9/49 (18.4%) patients with mild disease (*p* > .05)

Levels of IL-6, sIL-2rα (sCD25), IL-1β, IL-1Ra, IL-8, IL-17A, IL-18, IL-22, IFN, IL-10, and TNF-α were analyzed (Table 3). IL-6, IL-10, sIL-2rα, IL-1Ra, and IL-18 levels were significantly elevated on admission in patients who developed a critical disease. A cut-off value for each of these markers was determined to predict the risk of developing a critical COVID-19 (Figure 1). Values of IL-6 > 142.5 pg/mL, IL-10 > 10.8 pg/mL, IL1β > 4.68pg/ml, sIL-2rα > 804.5 pg/mL, IL-1Ra > 88.4 pg/mL, and IL-18 > 144 pg/mL were associated with the development of critical COVID-19 in the age-adjusted univariate analysis. In the age-adjusted multivariate analysis, this association was confirmed only for IL-10, sIL-2rα, and IL-18 (Table 4).

**Table 3.** Comparison of cytokine levels on admission in mild, severe, and critical COVID-19.

|  | **Mild**<br>**n = 41** | **Severe**<br>**n = 31** | **Critical**<br>**n = 38** | **P value** |
| --- | --- | --- | --- | --- |
| **IL-6** | 28.0 (13.0-76.0) | 36.0 (18.0-87.0) | 218.5 (47.6-819-7) | **P < .001** |
| **IL-10** | 0.0 (0.0-14.2) | 4.1 (0.0-36.5) | 63.1 (21.5-112.9) | **P < .001** |
| **sIL-2α** (sCD25) | 519 (364.2-768.0) | 701 (474.0-795.0) | 972 (579.0-1338.0) | **P = .012** |
| **IFN** | 5.1 (1.4-13.0) | 11.7 (1.6-39.7) | 13.9 (3.1-33-7) | n.s. |
| **IL1β** | 2.9 (0.9-2.0) | 7.9 (1.3-12-3) | 11.2 (5.1-18.5) | **P = .001** |
| **IL-1Ra** | 47.7 (24.6-96.5) | 54.8 (32.0-125.4) | 104.5 (49.3-333.0) | **P = .001** |
| **IL-8** | 42.8 (14.5-93.9) | 51.5 (20.5-116-2) | 49.7 (21.7-109.4) | n.s. |
| **IL-17A** | 0.0 (0.0-2.7) | 0.0 (0.0-3.8) | 0.0 (0.0-9.17) | n.s. |
| **IL-18** | 51.0 (17.9-92.4) | 46.8 (32.8-93.9) | 181.8 (87.3-307.5) | **p < .001** |
| **IL-22** | (0.0-0.0) | 0.0 (0.0-72.1) | 0.0 (0.0-0.0) | n.s. |
| **TNF** | 31.8 (19.4-54,9) | 37.7 (22.7-63.3) | 50 .7 (35.7-79.3) | n.s. |

Abbreviations: COVID-19, coronavirus disease 2019; IFN, interferon; IL-1Ra, IL-1 receptor antagonist; n.s., not significant; TNF, tumor necrosis factor; sIL-2 $\alpha$ , serum IL-2 receptor alpha.

**Table 4.** IL-6, IL-10, sIL-2rα (sCD25), ll1β, IL-1Ra, and IL-18 cut-off values and ORs (95% CI) for the risk of developing critical COVID-19 in the age-adjusted univariate and multivariate analyses.

| Variable | Cut-off<br>(pg/mL) | OR (95% CI) Univariate<br>model | OR (95% CI) Multivariate<br>model |
| --- | --- | --- | --- |
| **IL-6** | 142.5 | 8.76 (3.56-21.54) | -- |
| **IL-10** | 10.8 | 20.73 (5.83-73.71) | 8.09 (1.73-37.97) |
| **sIL-2α (sCD25)** | 804.5 | 12.46 (4.00-38.8) | 11.07 (2.39-51.32) |
| **IL1β** | 4.68 | 4.57 (1.81-11.53) | -- |
| **IL-1Ra** | 88.4 | 4,14 (1.82-9.41) | -- |
| **IL-18** | 144.0 | 16.10 (5.83-44.46) | 10.68 (2.87-39.69) |

Abbreviations: CI, confidence interval; COVID-19, coronavirus disease 2019; IL-1Ra, IL-1 receptor antagonist; OR, odds ratio; sIL-2 $\alpha$ , serum IL-2 receptor alpha (sCD25).

Lymphocyte subsets were analyzed in peripheral blood in a subgroup of patients on admission. The total number of T, B, and natural killer (NK) cells was significantly decreased in patients with more severe disease (Table 5). Neither a univariate nor a multivariate analysis was performed due to the small number of patients with critical COVID-19.

**Table 5.** Lymphocytes CD3, CD4, CD8, CD4/CD8 ratio, CD19, and NK cells in patients with mild, severe, and critical COVID-19.

|  | Mild (n = 39) | Severe (n = 23) | Critical (n = 14) | *P* value |
| --- | --- | --- | --- | --- |
| **CD3** | 1411<br>(986-2137) | 938<br>(639- 1426) | 507<br>(238-938) | *P* < .05 |
| **CD4** | 950<br>(653-1390) | 794<br>(463-1034) | 256.<br>(163-594) | *P* = .00 |
| **CD8** | 478<br>(317-685) | 327<br>(132-408) | 190<br>(63-253) | *P* = .00 |
| **CD19** | 256<br>(170-358) | 166<br>(84-218) | 93<br>(74-158) | *P* < .05 |
| **NK** | 170<br>(130-287) | 143<br>(72-320) | 65<br>(37-123) | *P* < .05 |
| **CD4/CD8** | 2.2<br>(1.5-2.9) | 2.1<br>(1.5-3.4) | 2.2<br>(1.0-2.8) | *P* > .05 |

Abbreviations: COVID-19, coronavirus disease 2019; NK, natural killer.

### Microbiological study

Oropharyngeal or nasopharyngeal swab samples obtained from 89 patients on the day of admission were tested for SARS-CoV-2 by RT-PCR either targeting E, N, and RdRP genes (72 patients) or E gene alone (17 patients). No significant differences in median Ct values according to symptom severity were observed (Table 6). No statistically significant differences between Ct values and number of days from symptom onset on admission or mortality were found either. Ct values >34 (associated with a low viral load) for the E gene on hospital admission were found in 16/89 (17.9%) patients; 2 had critical COVID-19 and 5 had severe COVID-19. Additionally, the N and RdPR genes were studied in 8/16 patients and a Ct value >34 was found in 3 cases. SARS-CoV-2 RT-PCR was performed in blood samples from 73 patients. All the results were interpreted as negative except in 3 cases that gave an indeterminate result (only a Ct >38 for the N gene was detected).

**Table 6.** E, N and RdRp median Ct values and IQRs according to symptom severity. IQR.

|  |  |  |  |
| --- | --- | --- | --- |
| **Mild** | 29.7 (22,2 – 35.3) | 31.9 (25.7- 36,2) | 25.1 ( 20.5-32.5) |
| **Severe** | 27.6 (19.4 – 33.1) | 28.0 (21.9-33,9) | 27.0 (20.9-34.1) |
| **Critical** | 26.4 ( 21.9-29.5) | 29.9 (25.5-31.2) | 26.9 (22.9-30.4) |

Abbreviations: Ct, cycle threshold; IQR, interquartile range.

## Discussion

Our study has identified a number of biomarkers which, in combination with the clinical classification, may be used to better evaluate the severity of COVID-19 and optimize therapeutic management strategies. Moreover, we propose cut-off values for each of these markers to predict the risk of developing critical COVID-19. sCD25, IL-1Ra, and IL-18 are of special interest as they have rarely been described as prognostic factors in COVID-19.

CRP, D-dimer, and ferritin have been described early in the pandemic as markers of hyperinflammatory state and disease severity [7, 16, 17], and COVID-19 guidelines have suggested that measuring their levels may have prognostic value [18]. Ferritin may be produced by activated pulmonary macrophages, and COVID-19 systemic inflammation is now considered as part of the spectrum of hyperferritinemic syndromes because a significant increase in ferritin levels has been found in severe compared with non-severe COVID-19 patients [19-22]. Similarly, CRP and D-dimer have been identified as prognostic markers, and cut-off values have been suggested in several studies: CRP > 4.96 mg/dL and D-dimer > 2600 ng/mL have been associated with critical illness in a study in China [7], whereas D-dimer > 500 ng/mL or >1000 ng/mL was found to be a significant risk factor for death in other studies [17, 23, 24]. However, in our study, the cut-off value of 440 ng/mL proposed for D-dimer was associated with a higher risk of developing critical disease in the univariate analysis but not in the multivariate analysis and neither was it associated with PE since D-dimer concentrations >440 ng/mL were observed on admission in only 9/15 patients who were later diagnosed with PE during hospitalization. NLR has already proven its prognostic value in cardiovascular and inflammatory diseases, in several types of cancer, and in some bacterial diseases [25]. Since the beginning of the COVID-19 pandemic, lymphopenia and increased NLR have been widely described as markers of severity [9, 16, 17]. Different mechanisms have been proposed for COVID-19-related lymphopenia: T cell exhaustion, apoptosis, pyroptosis, and a direct cytopathic effect of the virus [26]. Flow cytometric evaluation was useful in our study to investigate this lymphopenia. We found a significant reduction of all lymphocyte subsets —CD3+, CD4+, and CD8+ T cells (with no inversion in the CD4+/CD8+ ratio), NK cells, and B lymphocytes— in severe/critical patients compared to mild patients, in line with findings from other studies [16, 27, 28]. In addition, CD3+CD8+ T cell counts ≤75 cells/μL have been associated with death [23].

Immunological markers were investigated. Since hypocomplementemia has been reported in several viral infections and activation of the complement system and vascular deposition of complement components have been found in SARS-CoV thrombotic microangiopathy, the complement system was assessed. Neither our study nor a previous report found lower C3 or C4 levels in severe COVID-19 patients [29], but a relationship between PE and low C4 levels cannot be excluded. However, a recent study addressed the activation of the complement system measuring C3a, C3c, and the terminal complement complex and found significant higher levels in ICU COVID-19 patients compared to non-ICU COVID-19 patients [30].

Eleven cytokines or their receptors were tested on admission in the 120 patients, and IL-6, IL-10, IL1β, IL-1Ra, sIL-2rα (sCD25), and IL-18 were found to be of prognostic value.

High serum levels of IL-6 have been widely described as a hallmark of severity in COVID-19 [9, 11, 16, 31, 32]. IL-6 plays a central role in the cytokine storm as it induces cytotoxic T lymphocytes, promotes Th17 cell lineage, inhibits regulatory T cells, and activates B cells and antibody production. A recent systematic review and meta-analysis [33] concluded that threefold higher serum IL-6 levels were found in patients with severe COVID-19 compared with those with noncomplicated disease, and that such increased levels were significantly associated with adverse clinical outcomes, including ICU admission, acute respiratory distress syndrome, and death. Furthermore, several studies have suggested a benefit of IL-6 and IL-6 receptor antagonists for the management of critical COVID-19 patients [12, 21, 34].

We found that both IL-1Ra and IL-10 anti-inflammatory cytokines were significantly elevated in critical cases compared with moderate or severe ones. IL-10 has been described early in the pandemic as a marker of severe disease [9, 31]. IL-1Ra produced by activated macrophages is a competitive antagonist for IL-1 and controls inflammatory responses, modulating the production of other inflammatory cytokines. IL-1Ra has been previously investigated in several small studies and increased levels have been found in severe clinical cases [35, 36], supporting its role as an important marker of disease severity.

IL-18 is a proinflammatory cytokine that facilitates IFN-γ production by Th1 cells in conjunction with IL-12 and activation of CD8+ T cells. In our study, significantly higher IL-18 levels were found in critical patients compared with moderate or mildly ill patients. A correlation between IL-18 and the severity of COVID-19 had only been found in a small study [8], but a recent study has suggested this cytokine has a prognostic value [37].

The serum sIL-2rα (sCD25) level is considered an important disease marker in hemophagocytic syndromes/hemophagocytic lymphohistiocytosis [38]. However, its value as a prognostic factor in COVID-19 has been rarely reported. A small preliminary study in China with 21 patients found significant increased serum sCD25 levels in critical patients compared with patients with moderate disease [16], and the authors have suggested that sCD25 may act as a negative regulatory factor for T cells, contributing to lymphopenia. Other investigators have suggested the relevance of the sIL-2rα /lymphocyte index for early identification of severe COVID-19 and prediction of the clinical progression of the disease [39]. In our study, sCD25 was accurate in estimating the risk of complicated disease, as a cut-off value of > 804.5 pg/mL yielded a 73.3% sensitivity and 76.0% specificity for developing critical COVID-19.

Finally, virological markers were studied as prognostic factors for the development of severe disease, but our results failed to confirm any association. We did not find a correlation between Ct values determined by RT-PCR from nasopharyngeal swabs specimens and severity of the disease. Previous studies have reported conflicting results: a correlation between lower Ct values (representing higher viral loads) in respiratory samples and greater disease severity has been shown in certain studies [40-43], but other studies of non-SARS-CoV-2 respiratory viruses have not shown such correlation [14, 40-42, 44]. SARS-CoV-2 testing of blood samples from critical COVID-19 patients who died was negative in 70 patients and indeterminate in 3. The detection of SARS-CoV in plasma was associated with critical disease in the 2003 SARS pandemic [45], but COVID-19 studies have been scarce. One study detected SARS-CoV-2 viremia in up to 41% of patients admitted in a hospital in Zhejiang province, China [46].

Our study has some important limitations: it took place in a very specific geographic area (two hospitals in the Mediterranean island of Majorca), and the number of patients is small. In addition, patients were admitted at different times during the course of the COVID-19 pandemic and received different therapies, which may have altered the outcomes.

In conclusion, ferritin, D-dimer, CRP, NLR, and cytokines and cytokine receptors IL-6, IL-1Ra, sIL-2rα (sCD25), IL-18, and IL-10 contribute, in association with clinical data, to the early identification of patients at risk of critical COVID-19 and susceptible of a more intensive therapy. Further studies evaluating these biomarkers and their ability for predicting critical COVID-19 are needed.

## Data Availability

All data are available on request to the corresponding author.

## List of abbreviations

Ct values: Cycle threshold values
CRP: C-reactive protein
IL-1Ra: IL-1 receptor antagonist
IFN: interferon
NK: Natural killer cells
NLR: neutrophil/lymphocyte ratio
PE: pulmonary embolism
sIL-2rα or sCD25: soluble IL-2 receptor alpha
TNF-α: tumor necrosis factor alpha.

## Declarations

- The study was approved by the local Ethics Committee (Comité Ético de Investigación Clínica Illes Balears n° IB 4169/20 PI) and was performed in compliance with the Declaration of Helsinki. Informed consent forms were obtained from all participants.
- The authors have no conflicts of interest to declare.
- This work was supported by a grant from Instituto Salud Carlos III [grant number **COV20/00943]**

